# Assessing COVID-19 Risk, Vulnerability and Infection Prevalence in Communities

**DOI:** 10.1101/2020.05.03.20089839

**Authors:** Amin Kiaghadi, Hanadi S. Rifai, Winston Liaw

## Abstract

**Background:** The spread of coronavirus in the United States with nearly one million confirmed cases and over 53,000 deaths has strained public health and health care systems. While many have focused on clinical outcomes, less attention has been paid to vulnerability and risk of infection. In this study, we developed a planning tool that examines factors that affect vulnerability to COVID-19.

**Methods:** Across 46 variables, we defined five broad categories: 1) access to medical, 2) underlying health conditions, 3) environmental exposures, 4) vulnerability to natural disasters, and 5) sociodemographic, behavioral, and lifestyle factors. We also used reported rates for morbidity, hospitalization, and mortality in other regions to estimate risk at the county (Harris County) and census tract levels.

**Analysis:** A principal component analysis was undertaken to reduce the dimensions. Then, to identify vulnerable census tracts, we conducted rank-based exceedance and K-means cluster analyses.

**Results:** Our study showed a total of 722,357 (~17% of the County population) people, including 171,403 between the ages of 45-65 (~4% of County’s population), and 76,719 seniors (~2% of County population), are at a higher risk based on the aforementioned categories. The exceedance and K-means cluster analysis demonstrated that census tracts in the northeastern, eastern, southeastern and northwestern regions of the county are at highest risk. The results of age-based estimations of hospitalization rates showed the western part of the County might be in greater need of hospital beds. However, cross-referencing the vulnerability model with the estimation of potential hospitalized patients showed that part of the County has the least access to medical facilities.

**Conclusion:** Policy makers can use this planning tool to identify neighborhoods at high risk for becoming hot spots; efficiently match community resources with needs, and ensure that the most vulnerable have access to equipment, personnel, and medical interventions.

## Introduction

The outbreak of the novel Coronavirus was first reported in Wuhan, China but has since spread to almost every country in the world. The highest number of cases and deaths, as of this writing, has been reported in the U. S. [1] Within the 50 states, there is an apparent disparity in the number and causes of infections and their spread within each state. However, what is common to all cases reported thus far, is the rates of mortality and hospitalizations that appear to be highest among relatively older populations and populations with underlying medical conditions that facilitate morbidity due to COVID-19 [2–8].

Much research has focused on clinical outcomes, epidemiological modeling, and transmission dynamics of the novel coronavirus (see for example, [9–12]), but less focus has been placed on risk and vulnerability to contracting the disease. Emerging studies have begun to report on the impacts of social vulnerability on COVID-19 from an incidence and outcome standpoint [2–7,13]. However, the spatial resolution of most studies to date has been at the global or country level, and less attention has been paid to finer spatial resolutions such as the census tract scale within a county. A finer spatial resolution is important from a vulnerability and risk standpoint as demonstrated in a recent study that showed that the poorest neighborhoods in Houston, Texas, might be at a higher risk of hospitalization from COVID-19 [14] based on an analysis of the Centers for Disease Control (CDC) underlying risk factors for severe COVID-19 cases [4] that include: asthma, Chronic Obstructive Pulmonary Disease (COPD), heart disease, hypertension, diabetes, and a history of heart attacks or strokes.

While the aforementioned underlying medical conditions are important risk factors, they weigh in on the risk of hospitalization but not necessarily on the risk of contracting the disease. As such, underlying medical conditions and sociodemographic variables may not fully represent the magnitude of the risk and the challenge in managing and mitigating disease in affected populations from pandemics such as COVID-19. Environmental pollutants such as air quality [15], CO_2_ emissions [13], and ambient conditions such as temperature and humidity [5,16] showed correlations with COVID-19 morbidity. Furthermore, environmental exposures due to proximity to contaminated areas such as Superfund sites, hazardous waste sites, landfills, and leaky petroleum tanks has long-term adverse effects on public health, immune systems, and vulnerability to certain diseases [17–21]. Public health is further exacerbated by natural disasters, such as hurricanes and severe storms [22–24] that expose populations to pathogens and pollutants in floodwater and their flooded homes and potentially contribute to weakened immune systems. Behavioral and lifestyle factors could also affect the vulnerability of a population to an infectious disease such as COVID-19. Obesity, in recent COVID-19 data, has been shown to be prevalent in hospitalized patients [7], and smoking has been associated with disease progression [25]. Finally, it should be noted that because the risk is unevenly distributed, shortages in hospital beds, personal protective equipment (PPE), and medications have emerged in some but not all communities [26–28], thereby widening disparities and exposing systemic shortcomings [29]. Limited access to medical facilities, especially with less than fully-functional transportation systems combined with lack of insurance coverage, could worsen the impact of COVID-19 for people with less favorable sociodemographic metrics and people in rural regions. Thus, a more holistic view of the vulnerability of communities to COVID-19 that considers all of the aforementioned variables is needed to guide decision-makers in identifying the areas and populations in their jurisdictions that require specific resources, response, and mitigation actions.

In this study, we develop a rigorous planning tool at the census tract level that examines influential determinants of vulnerability to COVID-19 in 5 broad categories (with 46 variables) that include: 1) access to medical, 2) underlying medical conditions, 3) environmental exposures, 4) vulnerability to natural disasters and 5) sociodemographic, behavioral, and lifestyle factors. However, understanding the vulnerability of a population to COVID-19 is only one aspect of planning for such a pandemic. Other aspects include expected morbidities, mortalities, and hospitalization rates. Thus, the goals for developing the planning tool are to better understand medical access gaps and demands for hospitalization, identify parts of the county where more protective measures and response actions need to be put in place, and have a data-driven framework for estimating case numbers, hospitalizations, and deaths by census tract. Another goal is to have a better sense of the number of persons that may be affected broadly and more specifically as authorities lift or modify current policies such as the Stay Home Work Safe policy in place for Harris County and the City of Houston.

Such a planning tool is critical in order to mitigate the impact of COVID-19 and prepare for future pandemics. Using this tool, policymakers can identify neighborhoods with a higher potential for becoming the next hot spots, efficiently match community resources with community needs, and ensure that equipment, personnel, medications, and support are available to everyone, particularly the most vulnerable and those in greatest need. This strategy is essential to address historical trends that have preferentially delivered resources to those with means resulting in gaps in quality [30–32]. The planning framework developed in the study is readily transferable to other counties in the US and can be expanded to the state level for decision-making on a short-term or long-term basis towards improving the overall health of communities in each state.

## Methods

### Study Region

Harris County, located in the southeastern part of Texas (Fig 1), is the third-most populous county in the U.S., with more than 4.7 million residents [33]. While ranked number 2 in the nation in terms of Gross Domestic Product (GDP) growth, the County exhibits geospatial socioeconomic disparities among its population. The County is experiencing fewer cases, and lower rates of transmission relative to the rest of the U.S. Fig 2 shows the number of confirmed cases of COVID-19 in Harris County compared to New York County, for example. As shown in Fig 2, both the total number of confirmed cases and the slope of the spread are significantly higher in New York compared to Harris County. This is important to note because it potentially offers the County the opportunity for using the developed tool for improved long-term planning to respond to community health needs and disparities in response to COVID-19 and other pandemics or natural disasters.

**Fig 1.**
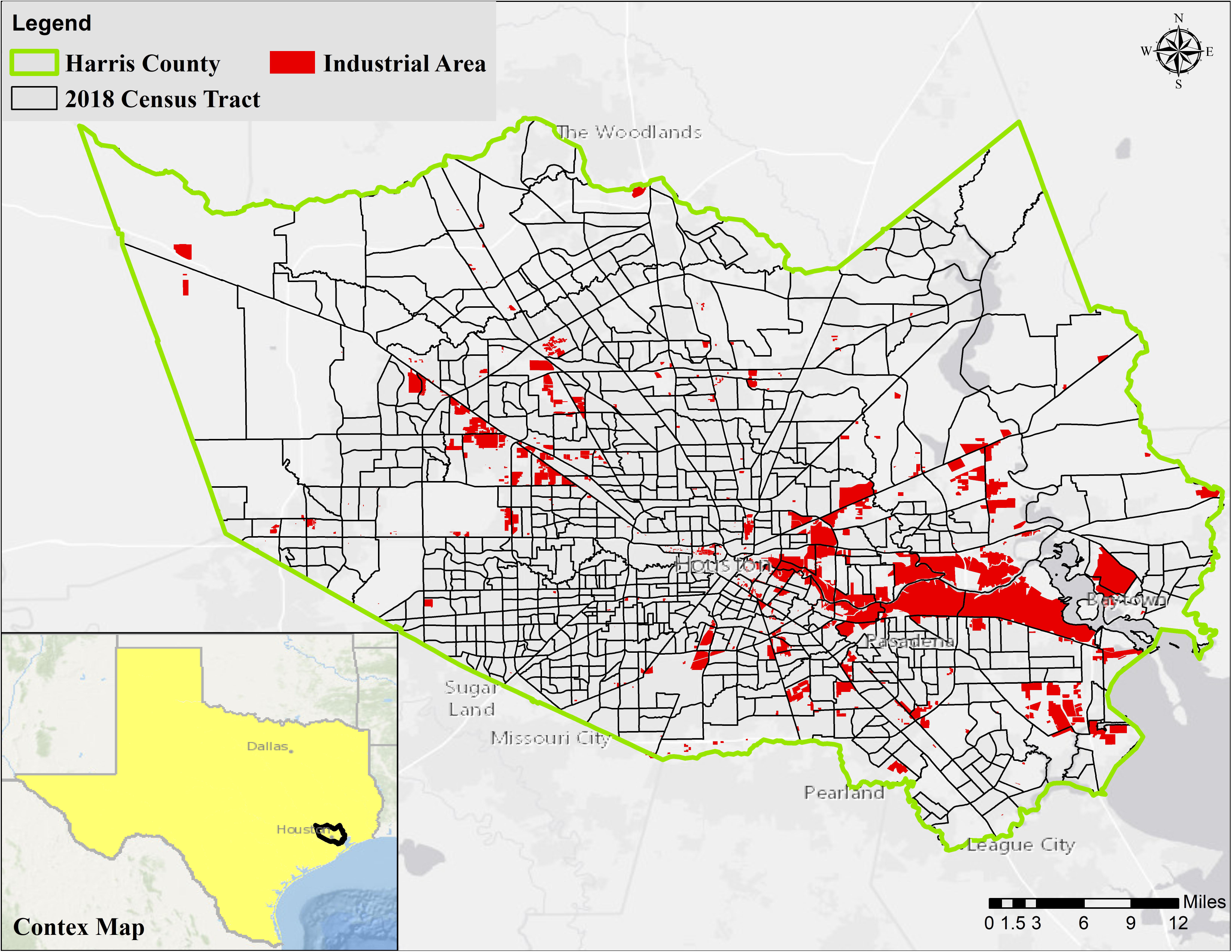
Map of Harris County in Texas and its 786 census tracts (2018). The industrial areas are defined according to the State of Texas classification of parcels.

**Fig 2.**
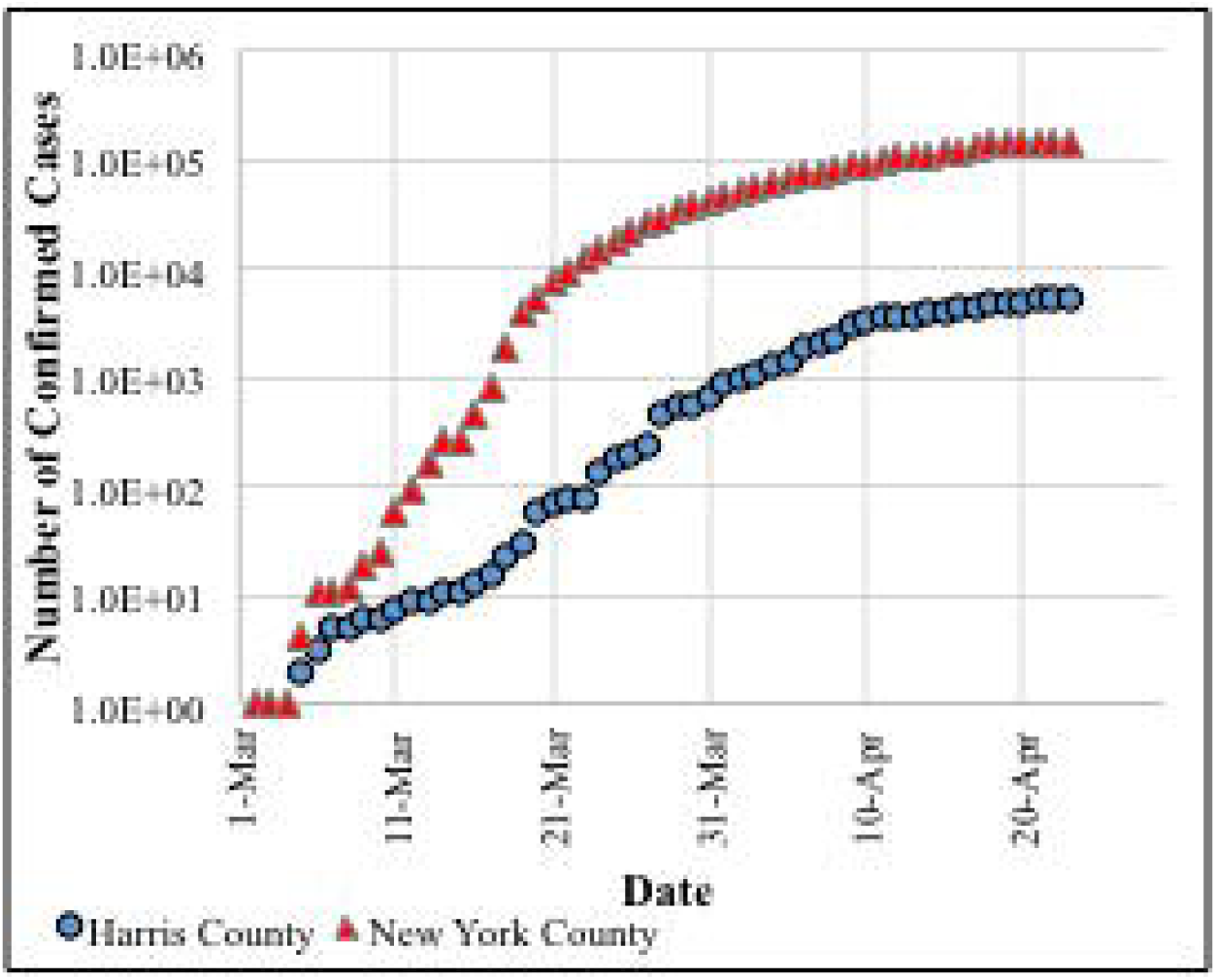
Number of confirmed cases of COVID-19 over time since March 1 in New York County and Harris County.

### Data Acquisition and Processing

All census data (2018) at the census tract level were compiled from the National Historical Geographic Information System (NHGIS) database [34]. Total population, the number of households, median income and income per capita (adjusted to 2018 US dollars), percent of the population below the poverty line, with cash public assistance or food stamps/SNAP (Supplemental Nutrition Assistance Program), living alone, with health insurance coverage, with a disability, education, and age distribution for each tract in Harris County were accessed. Using the detailed variables in the census data, education in this study was defined as the percent of the population with high school diplomas or higher degrees. Due to the importance of age in the vulnerability to COVID-19, both median age and the percent of the population in decadal age intervals were calculated. The percent of the population below the poverty line was chosen as the main economic variable, and the household density was calculated by dividing the total number of households by the area of each census tract.

Two measures of vulnerability to flooding were defined, using data from Hurricane Harvey that had severe impacts on Harris County in 2017: i) the ratio of the number of households that filed damage claims based on Federal Emergency Management Agency (FEMA) data [35] to the total number of houses in each tract, and ii) the ratio of the wetted areas (with water depth greater than zero) during Hurricane Harvey in a census tract to the total area of the tract (the specific methodology for this approach is described in [36]).

Locations and types of medical facilities including nursing facilities, federally qualified health centers, hospitals, rural health clinics, urgent care centers, and Harris County Health System facilities were obtained from the Health Resources and Services Administration (HRSA) query data explorer tool [37], Harris County Health System [38], and the Homeland Infrastructure Foundation-Level Data (HIFLD) database [39]. The Microsoft Bing Maps Platform APIs [40] was used to estimate the drive time from the centroid of each tract to all of the available medical facilities nearby. ArcMap was used to extract the coordinates of both origins (centroids) and destinations (medical facilities), and the minimum travel time in minutes was then recorded for each tract in Microsoft Excel.

The underlying conditions that might affect the vulnerability to COVID-19 (arthritis, asthma, high blood pressure (HBP), cancer (except skin cancer), high cholesterol, chronic kidney and heart diseases, COPD, diabetes, poor physical and mental health, and stroke); as well as increased-risk behaviors/conditions (binge drinking, smoking, no leisure time physical activity, obesity, sleep less than 7 hours), and preventive indicators (annual doctor and dentist checkups, medication for high blood pressure (HBP), cholesterol screening, and routine physical exams) were all acquired from the 500 cities mapper database [41] (Table 1), which draws from the Centers for Disease Control and Prevention’s (CDC) Behavioral Risk Factor Surveillance System. It should be noted that data from [41] were only available at 584 out of 786 (73.8%) census tracts in Harris County. Census tracts without data are clearly identified in all figures.

**Table 1.** Variables within each category (the choice of variables was based on the PCA analysis, previous studies, and data availability)

| Category | Name | Variables |
| --- | --- | --- |
| 1 | Access to Medical | Household density, drive time to a medical facility, access to HBP medications<sup>1</sup>, physical checkup<sup>1</sup>, dental checkup<sup>1</sup>, cholesterol screening<sup>1</sup>, insurance coverage<sup>1</sup>, routine physical exams<sup>1,2</sup> |
| 2 | Underlying Medical Conditions | Arthritis, Asthma, HBP, Cancer (except skin cancer), high cholesterol, chronic kidney disease, COPD, chronic heart disease, diabetes, poor mental condition, poor physical condition, stroke, at least one disability, median age, age above 50, age above 60, age above 70, age above 80 |
| 3 | Environmental Exposures | Distance to a hazardous site, number of hazardous pollution events and LPST<sup>3</sup>, number of dry cleaners, petroleum storage tanks, and IHWCA sites<sup>4</sup>, ozone concentration, NO<sub>2</sub> concentration, PM<sub>2.5</sub> concentration |
| 4 | Vulnerability to Natural Disasters | FEMA Harvey claims ratio, Harvey inundation ratio |
| 5 | Sociodemographic, Behavioral, and Lifestyle Factors | Binge drinking, current smoker, no physical activity, obesity<sup>5</sup>, low sleep quality, education beyond high school diploma<sup>1</sup>, below the poverty line, living alone, |

<sup>1</sup> The exceedance was calculated in the opposite direction.
<sup>2</sup> Routine physical exams include: Mammography (ages 50-74), Pap Smear use (ages 21-65), Fecal Occult blood test, Sigmoidoscopy, or Colonoscopy (ages 50-75), older men and women (+65) up to date on core clinical preventive services.
<sup>3</sup> Leaky petroleum storage tank (underground tank)
<sup>4</sup> Industrial and Hazardous Waste Corrective Action defined by the Texas Commission on Environmental Quality
<sup>5</sup> Obesity has emerged as a critical factor in hospitalization from COVID-19. In the context of this analysis, it was separated from other medical conditions in Category 2

As noted before, ambient conditions such as temperature and humidity could affect the spread of COVID-19; however, in this study, an ambient gradient in Harris County was neglected as the spatial change over the County is expected to be minimal. Three indicators of air quality: ozone, nitrogen dioxide (NO_2_), and particulate matter smaller than 2.5 micrometers (PM_2.5_), were downloaded from the Texas Air Monitoring Information System (TAMIS) database [42]. For ozone, the 8-hour average concentrations were calculated using IBM SPSS (version 26) for all of the available monitoring stations (40) and compared with the 70 ppb standard established by the United States Environmental Protection Agency (EPA). The number of exceedances of the EPA standard for each monitoring station over the period of 2000-2019 was then calculated. Interpolation tools in ArcMap were used to convert the median measured concentration for each station to a continuous raster to overcome the spatial sparsity in measurements. The generated raster was then used to calculate the concentration of ozone for each census tract using the zonal statistics tool in ArcMap. One particular station (695: UH Moody Tower) was removed from the ozone calculation due to its extremely low temporal resolution compared to the other stations. Similar approaches were taken for NO_2_ (hourly measurements for 21 stations with 100 ppm standard) and PM_2.5_ (averaged daily data for 12 stations with a 35 μg/m^3^ standard).

Environmental releases from various sources to air, water, soils in Harris County were obtained from the United States Coast Guard National Response Center database [43]. The total number of emissions, pollution spills, or contaminant discharge events that occurred during the period between 2000 and 2020 for each zip code was extracted from the database by combining different years, filtering the actual events for Harris County, and removing redundant data points. The spatial join tool in ArcMap was used to convert the total number of events in each zip code to a summed total number of events for each census tract. The resulting value was added to the number of leaking petroleum storage tanks (underground and aboveground tanks) reported by the Texas Commission on Environmental Quality (TCEQ) [44] in the tract. A hazardous sites shapefile was developed by merging two databases: the EPA Superfund Enterprise Management System database [45], and the Texas Commission on Environmental Quality (TCEQ) GIS database [44]. From the latter source, the locations of municipal solid waste sites/landfills were acquired. The Near tool in Arcmap was used to calculate the distance between the centroid of each census tract to the nearest aforementioned hazardous sites. A second environmental variable was defined as the sum of the total number of dry cleaners, petroleum storage tanks (all underground and aboveground tanks), and sites that are part of an Industrial and Hazardous Waste Corrective Action (IHWCA) program located within each census tract; data for those was obtained from [44]. Both Shapiro-Wilk and Kolmogorov-Smirnov tests conducted in IBM SPSS showed that none of the datasets were normally distributed.

### Defining Categories

A Principal Component Analysis (PCA) with orthogonal rotation (Varimax with Kaiser Normalization) was conducted in IBM SPSS as the first step to reduce the dimensions. Due to the limitation in data availability, as noted before, the PCA was performed for data from 584 tracts with all available data. Eigenvalues from random values were generated and compared with the values in this study using a parallel analysis engine [46]. This comparison was made to determine the number of components that should be retained in the analysis; components with eigenvalues greater than the randomized method were kept. The first five components that could explain ~ 80% of the variability in the 46 variables showed eigenvalues larger than the ones generated by the engine. S1 Table in the Supplementary Information (SI) shows the most dominant variables in each component (category).

The choice of variables for the study (Table 1) was based on the results of the PCA in addition to findings reported in previous studies, and data availability. Category 1 includes access to medical care, including medical facilities, medications, and insurance coverage, routine checkups, and physical exams, as well as household density as a surrogate for interaction among individuals within each tract (e.g., how crowded grocery stores could be in the tract). Category 2 includes chronic diseases, medical conditions, disability that could potentially affect the vulnerability to COVID-19, and age distribution. For environmental exposure, pollution events from various sources, the 3-air quality indicators, and the presence of hazardous sites were included. Flooding from Hurricane Harvey was the only metric in Category 4, although this could be expanded in future work to include heat, drought, wildfires, and other natural disasters. Finally and for Category 5, a combination of social, economic, behavioral, and lifestyle factors that could potentially threaten the health of individuals during the COVID-19 pandemic was considered.

## Statistical Analyses

### Vulnerability Analysis

Two classification approaches were used in the study with the goal of identifying the most vulnerable populations to COVID-19; a rank-based exceedance method developed in Microsoft Excel, and a standard K-Means Cluster Analysis (K=3) using IBM SPSS. Validation of any developed models for the vulnerability was not possible due to lack of data at the desired spatial resolution and the fact that the pandemic is still developing. Thus, the second model (K-means) was used as a benchmark for the first model for comparison purposes.

In the rank-based exceedance method, for each variable, sorting the data in Microsoft Excel developed the rank of each census tract relative to other tracts within Harris County. The exceedance rate (percentile) was calculated as follows:

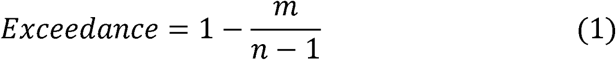

Where m is the rank, and n is the total number of tracts (786 in Harris County). The calculated exceedance for a given tract represents the percent of tracts that have a better condition than the selected one. To ensure that the direction of exceedance is the same among all variables, (1 – exceedance) was used for variables with positive nature such as insurance coverage, education, access to medication, and preventive tests. For each category, the average value of exceedance for all of the variables within that category was calculated and reported. In addition to classifying the tracts for each of the aforementioned categories, an overall vulnerability was defined by averaging the exceedance rates of the five defined categories. The percentile associated with each averaged value (for each category and for the overall vulnerability) was calculated and exported to ArcMap to generate decision-support level maps.

In the K-means cluster analysis (K-means is an unsupervised machine learning algorithm), three classes were defined for each category. As a result, the output classes were ordered as high (severe), average, and low depending on the order of the final cluster centers. The ANOVA test was conducted on the clusters to ensure that the values of the different variables were significantly different between clusters. Similar to the exceedance method, an overall vulnerability for each census tract was determined by averaging the five output class numbers (i.e., 1, 2, and 3) associated with the five categories. For illustration purposes, the percentile rank for each of the tracts was calculated and exported to ArcMap.

Although it is possible to assign weights to the categories and calculate a weighted average, equal importance for the categories was assumed in this study. Assigning weights is beyond the scope of this paper as there is not enough evidence to support such assignments as of this writing.

### Morbidity, Hospitalization, and Mortality Rates

As of April 23^rd^, 2020, the total number of confirmed cases and deaths in Harris County were 5,330 and 82 [47], respectively, which translates to a morbidity rate of 121.83 per 100,000 persons, and 1.54% mortality rate. The COVID-19 hospitalization rates for the two Trauma Service areas Q and R (comprising the southeast region of Texas) are 7.86 and 9.84 per 100,000 persons [48]. It is noted that predicting the risk of infection, morbidity, and mortality rates in Harris County requires a more extensive dataset and would involve considerable uncertainty due to limited knowledge of the SARS-CoV-2 virus and the relatively limited number of tests that have been administered thus far. In this study, the distribution of COVID-19 by sex and age was the only metric considered for predicting the risk of hospitalization, morbidity, and mortality in Harris County. This is mainly due to limited access to health information for individuals in the County, such as comorbidities and the unknown effect of underlying medical or other lifestyle conditions on the risk of infection and potential complications from COVID-19.

According to the data obtained from the New York Department of Health and Mental Hygiene [49], the morbidity rates (using the reported number of cases) observed in New York City (NYC) were 171.46, 1554.45, 2529.03, 2552.64, 2976.74, and 1687.8 per 100,000 population for age groups of 0-17, 18-44, 45-64, 65-74, and +75 years, respectively. Although the nature of the spread of COVID-19 could be substantially different in New York City compared to Harris County, the NY City rates were applied in this study to develop a worst-case scenario estimate. Age data from [34] were re-classified to align with the intervals defined by the New York Department of Health and Mental Hygiene [49].

As of April 11^th^, 2020, the hospitalization rates among people with laboratory-confirmed COVID-19 in the U.S., based on data from the CDC [50], were 1.1, 0.3, 10, 32.8, 45.8, 76, and 110 per 100,000 population for age groups of 0-4, 5-17, 18-49, 50-64, 65-74, 75-85, and +85 years, respectively. However, substantially higher rates were reported in New York City [49]; 13.55, 153.69, 630.34, 1192.5, 1830.07, and 437.24 per 100,000 population for age groups of 0-17, 18-44, 45-64, 65-74, and +75 years, respectively. For the CDC rates, the population with age under five years old was used instead of 0-4 years.

Three separate studies, one with 3,665 cases in mainland China and 1,334 cases detected outside of mainland China [51], another one with 73,780 cases in Italy [52] and the latest with 141,754 cases in New York [49] reported different mortality rates among different age intervals. Both studies in China and Italy reported the mortality rates among confirmed COVID-19 cases while the New York study reported the total of deaths per 100,000 persons within each age interval. In this study, rates from all three studies, as shown in Table 2, were used to calculate the mortality rates associated with COVID-19 in Harris County. Multiplying the rates in each study by the associated number and percentages of the population in each age group was used to calculate the risk of morbidity, hospitalization, and mortality rates for each census tract.

**Table 2.**
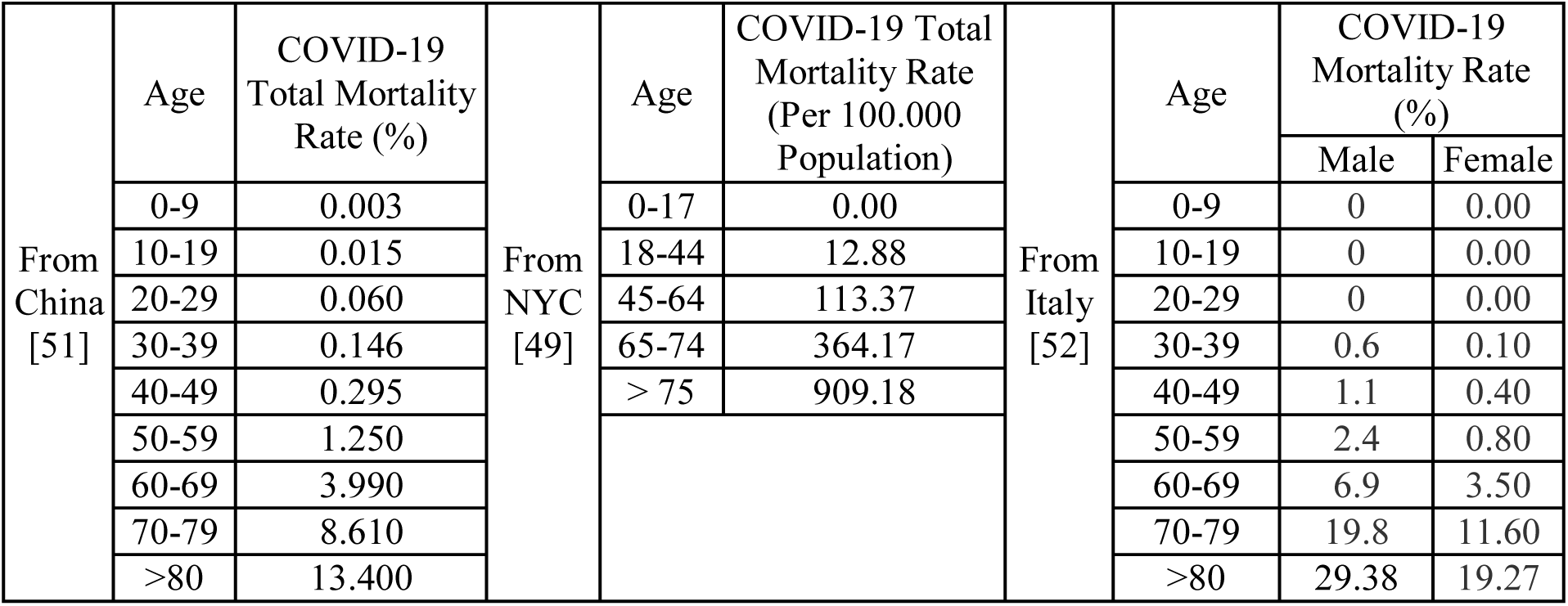
The mortality rate among COVID-19 confirmed cases by age only [51] by age and sex [52], and per 100,000 population [49].

An important caveat of the approach used in this study is the emerging realization of underreported positive cases in the US and potentially undercounting deaths by not testing all persons who have died in the US since December 2019. A second important caveat is that the true rate of infection is currently unknown. The third caveat is that Texas, as of this writing, has had one of the lowest rates of testing in the US.

## Results and Discussion

### Geospatial Distribution of Determinants in the 5 Categories

S2 Table provides a summary of statistics for all of the 46 variables used in the study. Among the 46 variables, maps are only presented for those that were not based on publicly available data. Fig 3 shows the locations of medical facilities (all types as described in Methods) within and around Harris County as well as the drive time to the nearest facility for each census tract. The drive time varies from seconds to 25.23 minutes, with a median of 4.74 minutes (S2 Table). As can be seen from Fig 3, people who live in areas located farther away from the center of the County, especially in the western and northeastern parts, have a longer drive time to a medical care facility. This longer drive time becomes even more critical if an individual does not have a personal car and needs to use the less than a fully functional transportation system. The travel time is even longer to facilities managed by the Harris Health system (typically used by individuals with no insurance or documentation). From a planning standpoint, Fig 3 below, when combined with vulnerabilities, can be used to drive decisions related to the establishment of field hospitals during periods of widespread transmission. Importantly, the data can be used to develop a more holistic response plan directing persons with various severity or symptoms of the disease to different types of medical intervention facilities.

**Fig 3.**
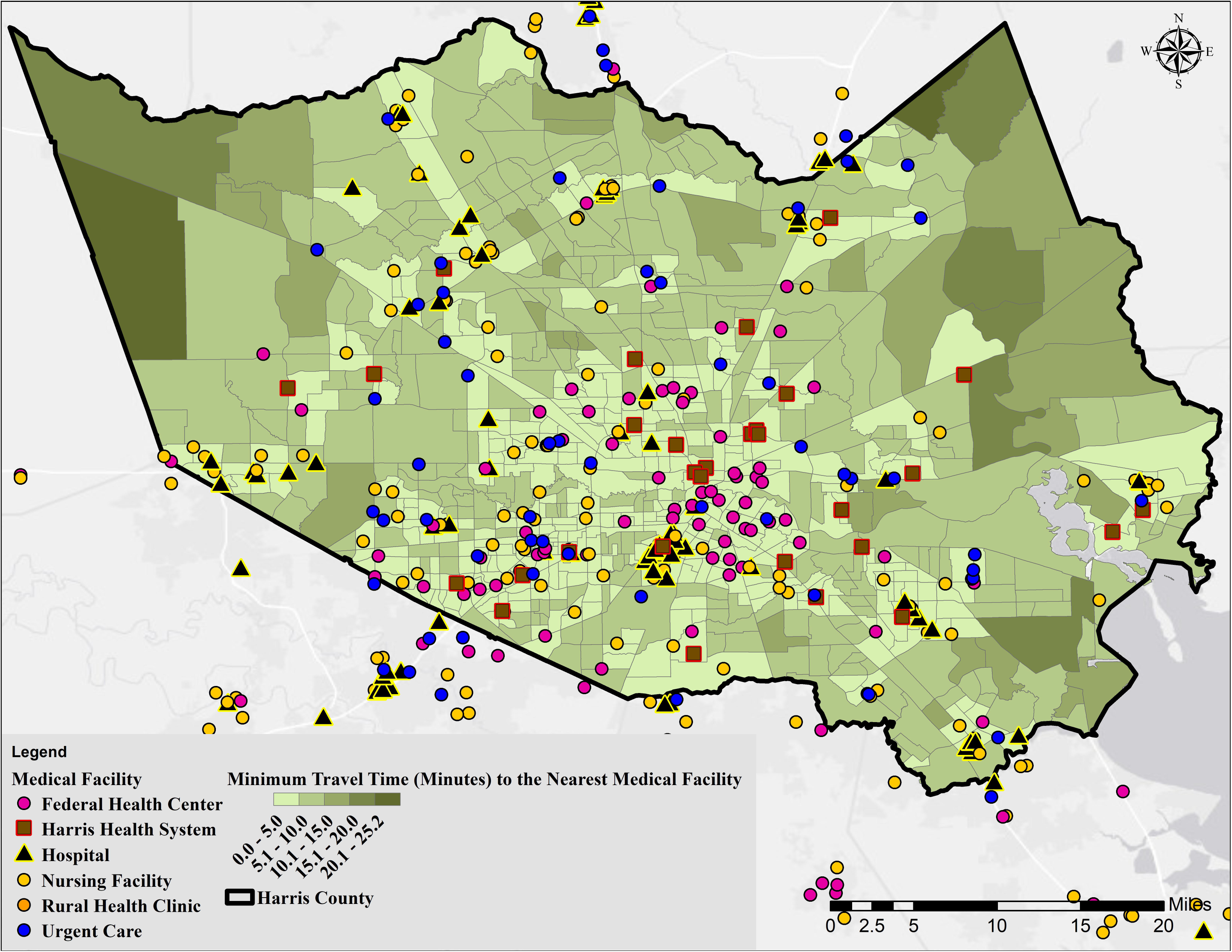
Map showing the distance from the centroid of census tracts to the nearest medical facility.

The distance to and location of hazardous sites (Superfund sites, landfills, and industrial hazards) are shown in Fig 4. The distance ranges from 79 to 9,386 m with a median of 2,105 m. The hazardous sites are spread over the entire County but are more concentrated closer to the industrial areas (see Fig 1), water bodies (Houston Ship Channel (HSC), and Galveston Bay (GB)). The tracts in less developed areas have the longest distance to a hazardous site indicating a higher potential vulnerability to pollution in the more developed parts of Harris County.

**Fig 4.**
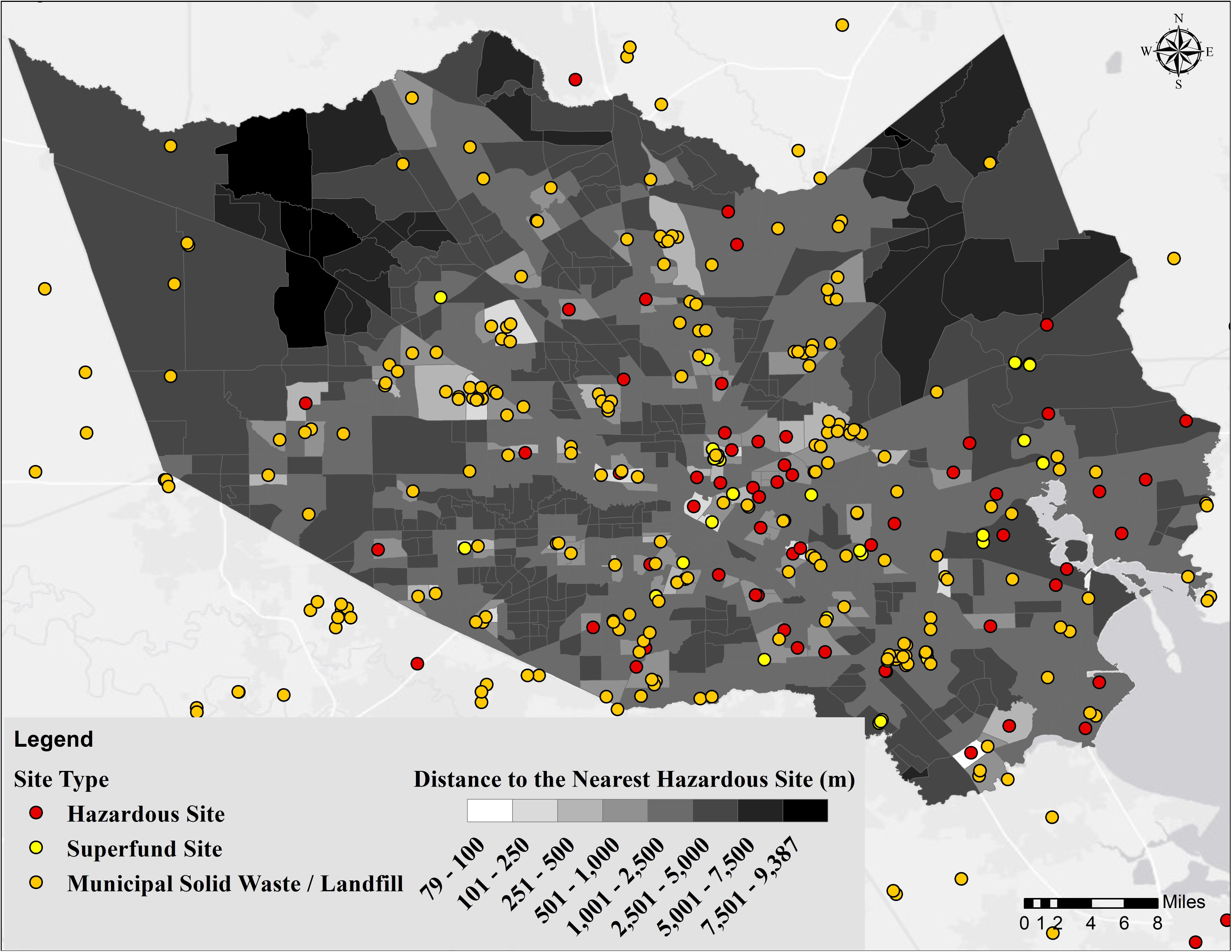
Map showing the distance from the centroid of census tracts to the nearest hazardous site.

S1-S3 Figs show the median concentration of ozone, NO_2_, and PM_2.5_ for each tract in addition to the number of times during 2000-2019 that a monitoring station exceeded the EPA standard. It should be noted that the number of stations and, consequently, the geospatial coverage was significantly lower for NO_2_ and PM_2.5_ when compared to ozone. Stations with the highest number of exceedances of EPA standards for all three measures are located near industrial areas (see Fig 1). While the central parts of the County showed the highest concentration of NO_2_ and PM_2.5_, ozone concentrations were highest closer to the industrial areas. The higher levels of NO_2_ in central parts of Harris County could be attributed to emissions from mobile sources that are more abundant in downtown Houston [53]. The observed pattern for ozone is a result of industrial activities, the ozone-NO_2_ relationship, and the wind pattern in Houston [54,55]. In the case of PM_2.5_, the higher concentrations in Harris County have been associated with regional aerosols, biomass burning, and gasoline combustion [56] that are higher in the central part of the County. The median concentrations for ozone, NO_2_, and PM_2.5_ were 21.24 ppb, 8.32 ppm, and 9.98 μg/m^3^, respectively, for the period of 2000-2019. What is interesting to note is the fact that the three variables have different spatial distributions thereby indicating potentially more important involvement in COVID-19 based on recent research showing increased vulnerability due to PM_2.5_ pollution in COVID-19 patients [57] and CDC’s indication that “people with moderate to severe asthma may be at higher risk of getting very sick from COVID-19.”

Contaminant discharge events (S4 Fig) and the second environmental variable representing dry cleaners, petroleum storage tanks, and IHWCA sites (S5 Fig) were substantially higher in industrial areas close to the HSC and GB: La Porte, Baytown, Deer Park, and Channelview, with the number of events as high as 1,449 (2000-present). The median was 12 events across all census tracts. Fig 5 shows the percentile of flooding across Harris County based on the filed claims to FEMA. Areas closer to the bayous/streams and flood control dams showed a higher vulnerability. A similar distribution was observed using the geospatial inundation modeling approach (S6 Fig). Finally, S7 Fig shows the distribution of educated persons in Harris County, defined as the ratio of age 25 years and over with a high school diploma or higher degree to the total population for each census tract.

**Fig 5.**
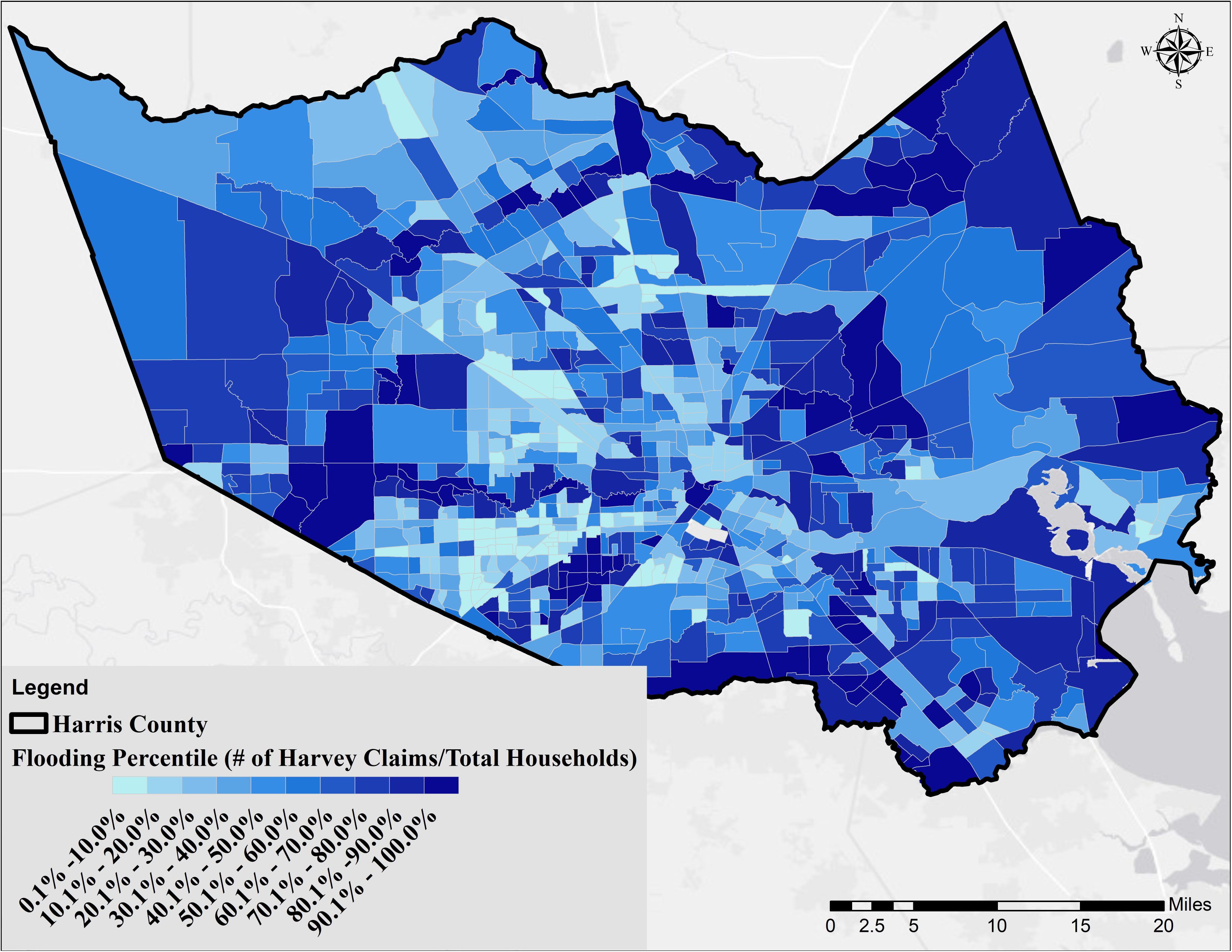
Flooding vulnerability based on the number of households that filed damage claims to the Federal Emergency Management Agency after Hurricane Harvey.

### Vulnerability in Census Tracts

Fig 6 represents the average exceedance for variables within Category 1 through Category 5 (values are reported in percentiles for the purpose of comparison among tracts), respectively, and Fig 7 shows the overall vulnerability for all variables in the five categories. Overall, the vulnerabilities associated with each Category exhibited varying geospatial distributions with some commonality (i.e., some census tracks had elevated vulnerabilities in each Category). Category 1, 2, and 5 vulnerabilities shown in Fig 6 (access to medical, underlying medical conditions, and sociodemographic, respectively) indicated a similar finding; the most severe vulnerability can be observed in areas with the least favorable conditions represented by the three categories (lowest income, lower education levels, less insurance coverage, unhealthy diet and lifestyles, and more underlying medical conditions). Category 3 (Vulnerability to Environmental) showed a spatially declining gradient from east to west with some hotspots around downtown Houston. This gradient could be explained by the presence of the majority of industrial activities in the eastern part of Harris County, and worse air quality near downtown Houston. Category 4 (vulnerability to natural disasters) showed the highest risk in the vicinity of major bayous/streams in the County, as discussed before.

**Fig 6.**
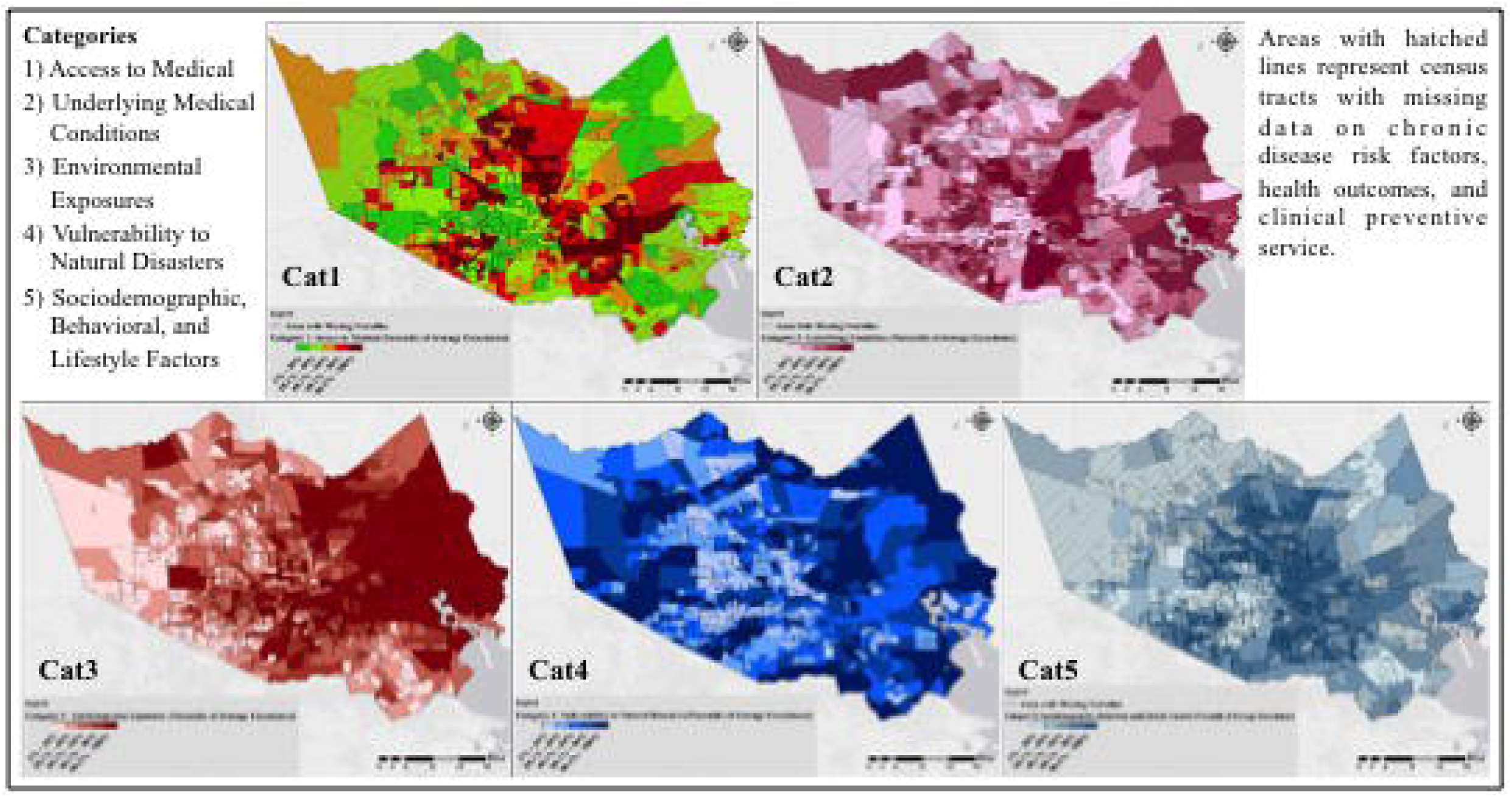
The average exceedance for variables in the 5 Categories. Averaged exceedance values are reported in percentiles for the purpose of comparison among tracts.

**Fig 7.**
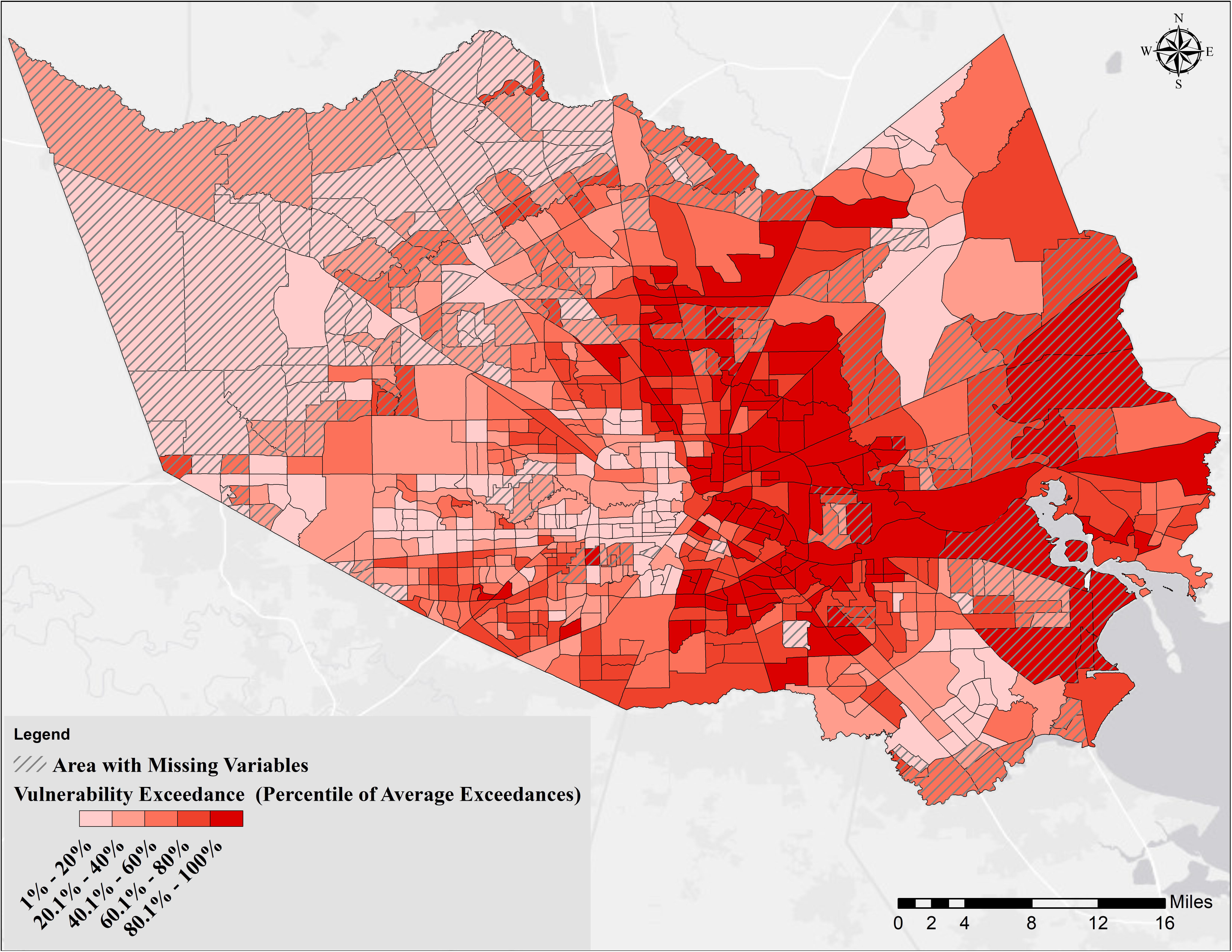
Overall vulnerability based on determinants in all 5 categories. Areas with hatched lines represent census tracts with missing data on chronic disease risk factors, health outcomes, and clinical preventive services [41].

Looking at Fig 7, it could be concluded that the most vulnerable persons to COVID-19 in Harris County, are living in the eastern part of the County, specifically areas next to the HSC and GB, and areas identified as opportunity zones [58]. The residents in these neighborhoods are individuals with the least favorable sociodemographics, are exposed to several chemicals (with industrial sources), and subject to flooding both from rainfall and storm surge (such as what was experienced during Hurricane Ike in 2008). Individuals living in the western and southeastern fringe of the County are least vulnerable. However, it is noted that the underlying medical condition data were not available for those tracts. It is also noted that individuals in those areas, if infected, especially in the western fringe, will have significantly limited access to medical facilities compared to the other parts of the County.

The developed tool can be used to look at the tracts individually and compare their vulnerability with other tracts. A spider chart, for example, enables users to look at all five categories at the same time and determine which category has the most influence on vulnerability within a given tract; S8 Fig shows the status of the tracts with the highest and lowest overall vulnerability in each category.

The final cluster centers for different variables for each category (K-means method) are represented in the S3 Table. S9-S13 Figs show the class of each tract (i.e., high/severe, average, and low) for Category 1 through Category 5, respectively. The results in all categories were similar to the exceedance methods, validating the choice of methodology. The overall vulnerability generated by the K-means methods led to a very similar map (S14 Fig) to the exceedance approach (Fig 7).

### Vulnerable Population Estimates in Harris County

For each category, the total population and the distribution of population in two age intervals, 45-65 (the age group with the highest number of reported COVID-19 cases), and +65 (the age group with the highest mortality rate), over different percentiles (from low to high with regards to the severity of conditions within each category) is shown in Table 3. Using the vulnerability findings presented above for Harris County (Fig 6, and yellow highlighted values in Table 3); a total of 59,307, 98,702, 78,723, 105,431, and 59,624 seniors (+65 years), who are at most risk of COVID-19 mortality, are living in areas with the highest vulnerability in Category 1 through 5, respectively. Considering the fact that Harris County is prone to flooding and the hurricane season is in progress from May through the end of November, a potential hurricane combined with the COVID-19 pandemic could lead to a compound natural disaster event affecting significant numbers of senior citizens as shown in Table 3. Decision-makers, to prepare for the worst-case pandemic scenario and occurrence of a hurricane, in particular, could use the numbers in Category 1 and 4 for planning response and recovery measures that take into account flooding and increased vulnerability to COVID-19. For overall vulnerability (Fig 7 and cyan highlights in Table 3), a total of 722,357 persons (~17% of the population of the County) including 171,403 with ages between 45-65 (~4% of the total population of Harris County), and 76,719 seniors (~2% of the population of the County), are at a higher overall risk.

**Table 3.** Distribution of the total population, and those of ages between 45 and 65, and above 65 years within different determinant category percentiles of vulnerability in Harris County.

| Percentile |  | 0% - 20% | 20% - 40% | 40% - 60% | 60% - 80% | 80% - 100% |
| --- | --- | --- | --- | --- | --- | --- |
| Category 1 | Total Population | 760,572 | 929,535 | 946,254 | 892,448 | 846,173 |
|  | 45-65 years | 212,214 | 236,982 | 232,755 | 210,859 | 173,524 |
|  | > 65 years | 113,479 | 103,052 | 96,236 | 80,253 | 59,307 |
| Category 2 | Total Population | 1,107,588 | 923,925 | 922,086 | 786,298 | 635,085 |
|  | 45-65 years | 234,744 | 222,531 | 230,052 | 204,197 | 174,810 |
|  | > 65 years | 71,517 | 86,353 | 98,296 | 97,459 | 98,702 |
| Category 3 | Total Population | 1,016,883 | 914,746 | 830,344 | 795,215 | 817,794 |
|  | 45-65 years | 269,504 | 224,015 | 202,769 | 182,543 | 187,503 |
|  | > 65 years | 108,117 | 95,234 | 90,642 | 79,611 | 78,723 |
| Category 4 | Total Population | 745,835 | 831,232 | 970,245 | 928,682 | 898,988 |
|  | 45-65 years | 163,690 | 199,625 | 235,323 | 230,867 | 236,829 |
|  | > 65 years | 68,795 | 86,866 | 95,116 | 96,119 | 105,431 |
| Category 5 | Total Population | 1,057,398 | 867,297 | 909,292 | 824,148 | 716,847 |
|  | 45-65 years | 292,819 | 220,506 | 212,651 | 188,267 | 152,091 |
|  | > 65 years | 123,228 | 101,213 | 93,823 | 74,439 | 59,624 |
| Overall<br>Vulnerability | Total Population | 998,996 | 927,584 | 906,212 | 819,833 | 722,357 |
|  | 45-65 years | 249,385 | 241,270 | 213,158 | 191,118 | 171,403 |
|  | > 65 years | 97,587 | 108,035 | 88,496 | 81,490 | 76,719 |

See Fig 6 for Categories 1 through 5 and Fig 7 for Overall Vulnerability geospatial distributions

### Vulnerability Association with Morbidity, Hospitalization, and Mortality

Fig 8 presents the geospatial distribution of averaged vulnerability across Categories 1 through 5, morbidity, mortality, and death estimates. The calculated morbidity rates, using NYC estimates [49], across the County, are shown in Fig 8B. This worst-case scenario would lead to a total estimated number of 70,018 COVID-19 cases with a morbidity rate of ~ 552 cases per 100,000 population (this is substantially higher than the reported cases to date of 5,330 as of April 23^rd^, 2020 [48]). There are 452,327 senior residents living in Harris County (10.3% of the total population). Among the seniors, for the NYC worst-case scenario, a total of 12,267 (2.71% of seniors and 0.28% of the total population) could be infected with COVID-19. By comparing the morbidity (age-based) and vulnerability results, as shown in Fig 8A and B, it could be concluded that ~ 10.0% of the total population and ~16% of seniors at the highest risk level (80%-100% percentile overall vulnerability) could contract COVID-19. While the actual morbidity rates in Harris County, to date, are lower than New York, China, and Italy, the specific reasons for this are unknown. This could be due to lower population density, relatively higher temperatures, or the fact that New York, China, and Italy had earlier reported and imported cases than Harris County, and social distancing was not deployed immediately or soon after. As mentioned previously, one caveat that places greater uncertainty for Harris County reported rates is the fact that Texas is listed as the 46^th^ state in terms of rates of testing (~542 per 100,000 population as of this writing [47]).

**Fig 8.**
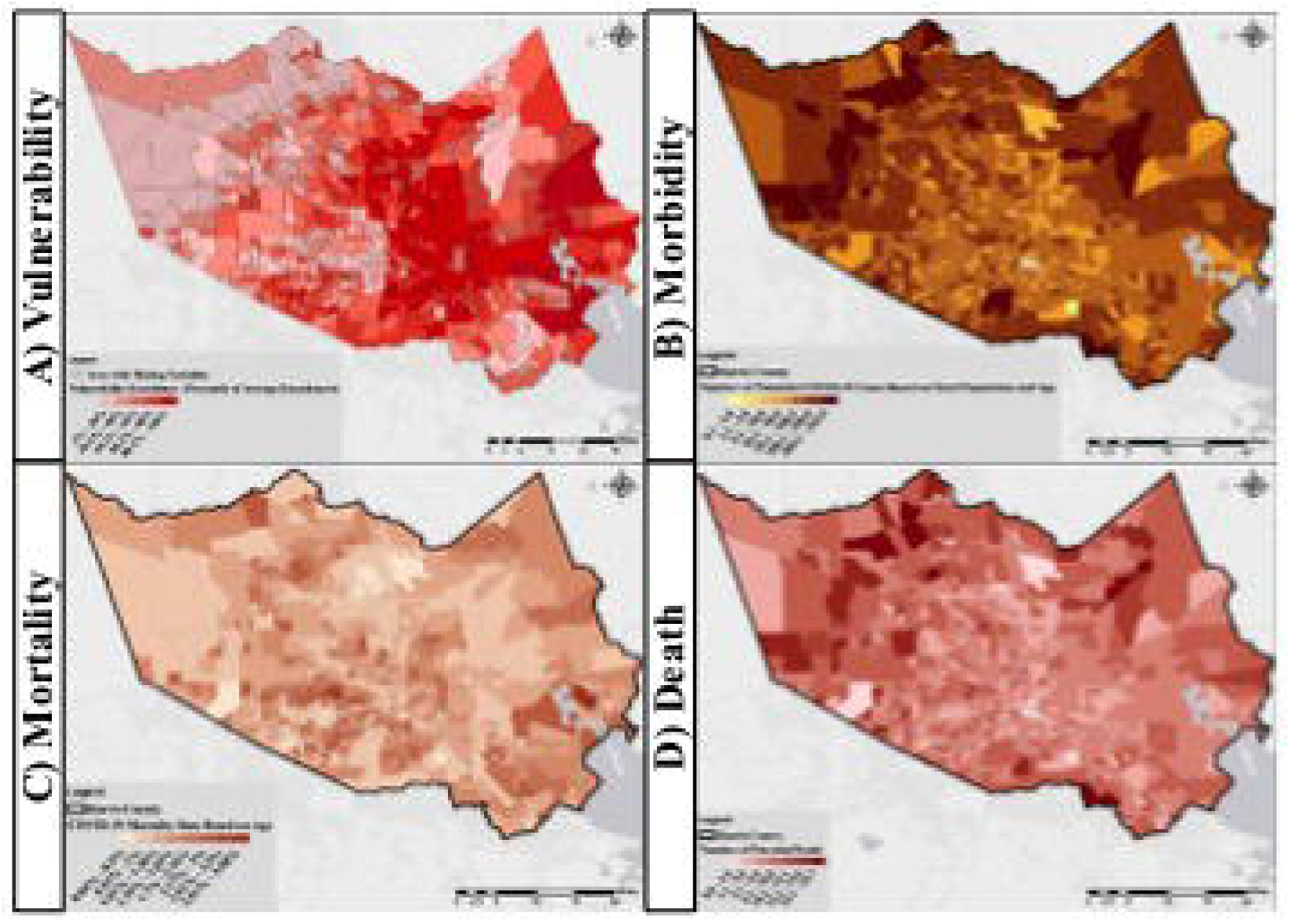
A) Overall vulnerability based on determinants in all 5 categories (see Fig 7), B) Morbidity rates in Harris County census tracts based on a worst-case scenario using NYC data [49], C) The mortality rate for each census tract solely based on the age distribution (rates associated with each age interval were extracted from [51]), and D) The number of potential deaths for each census tract in Harris County (the calculation was based on the age distribution within each tract and rates reported by the New York Department of Health and Mental Hygiene [49])

Estimating hospitalizations based on age distributions and hospitalization rates reported by CDC [50] resulted in a total of 672 hospitalized patients (S15 Fig) and a rate of 15.37 per 100,000 population over the County (this estimate is also in excess of the reported data for Harris County on a daily basis that is between 7.86 and 9.84 per 100,000 persons (obtained by dividing the current day number of hospitalized patients to the total population, [48]). A worst-case scenario using NYC data leads to 16,200 hospitalizations and a rate of ~366 per 100,000 persons (S16 Fig). As shown in S16 Fig, the highest rate of hospitalization is predicted for the northeast and northwest parts of Harris County, where people have the least access to medical facilities (Fig 3) and are prone to flooding (Fig 5).

Fig 8C (based on [51]) and S17 Fig (based on [52]) show the calculated mortality rates in Harris County census tracts. The average mortality rates (ratio of the number of total deaths to the number of confirmed COVID-19 cases) in Harris County, calculated based on the rates from China [51] and Italy [52] were 1.39% and 1.28%, respectively, which are both lower than the current rate of 1.54% in the County. Fig 8D shows the total deaths estimated in individual tracts in Harris County based on the rates reported by the New York Department of Health and Mental Hygiene [49]. A total of 4,020 deaths with a rate of 92.65 per 100,000 persons were estimated based on the age distribution (this is higher than the reported 82 deaths in Harris County as of April 23^rd^, 2020).

## Limitations

A number of limitations are noted that should be considered when interpreting these results. First, data was lacking for some of the variables for a number of census tracts in Harris County; the CDC’s 500 Cities dataset [41] is such an example. The analysis findings might change if a Houston-specific dataset is used. However, the downside is that the approach would not incorporate County level considerations. Second, and most important (this generally is true of numerous other COVID-19 studies), granular sub-county data about COVID-19 cases, hospitalizations, and deaths are lacking (at the zip code or census tract levels). It would be essential to have such granularity to validate/compare projections based on determinants against actual morbidities and mortalities. Using morbidity, hospitalization, and mortality rates reported in other regions places undue levels of uncertainty in the findings that would be substantially reduced when such data are available, or a national surveillance system [60] is in place.

## Conclusions

In this novel project, we develop a planning tool that can help identify populations at higher risk of infection, morbidity, and mortality from COVID-19 at the census tract level. These findings can guide the allocation of scarce resources, and thus, are relevant to policymakers at all levels of government. Effectively using the results from the planning tool to inform actions could mean the difference between suppressing the virus and allowing it to re-emerge. Studies, for example, would be needed to validate projections and findings. However, to do so, sub-county incidence, hospitalization, and mortality data describing COVID-19 cases in Harris County would be needed. In comparison, it is noted that studies that map the geospatial spread of coronavirus from Wuhan to neighboring communities are starting to emerge [59,60], and similar efforts need to be launched in the US. The application of geospatial methods to case data, enables significantly more rigor in understanding the confluence of various factors that jointly increase vulnerabilities and reduce resilience to COVID-19 spread, impact, re-emergence in new hot spots or on a seasonal basis. While geospatial indices exist [61,62], they are not tailored to the unique features of this virus. Lastly, the findings from geospatial analyses such as this study enable public health departments to efficiently and equitably allocate resources such as testing, contact tracing, and medical interventions to areas in greatest need.

## Data Availability

All data included in the manuscript.

## Competing Interests

The authors have no competing interests.

## Acknowledgments

This research was funded by the Hurricane Resilience Research Institute (HuRRI), an interdisciplinary research institute at the University of Houston focused on resilience to natural disasters and RAPID grant # 1759440 and grant # 1840607 from the National Science Foundation. Their support is gratefully acknowledged.

## Supporting information

**S1 Fig. Map showing the median of averaged 8-hr concentration of Ozone (ppb) in Harris County. The yellow dots show the location of air quality monitoring stations maintained by the TCEQ. The magnitude of the dots shows how many times the averaged 8-hr ozone concentration in a specific station exceeded the 70 ppb standard established by EPA from 2000-2019**

**S2 Fig. Map showing the median of hourly concentration of NO_2_ in Harris County. The yellow dots show the location of air quality monitoring stations maintained by TCEQ. The magnitude of the dots shows how many times the hourly NO_2_ concentration in a specific station exceeded the 100 ppb standard established by EPA from 2000-2019**

**S3 Fig. Map showing the median of averaged daily concentration of PM_2.5_ in Harris County. The yellow dots show the location of air quality monitoring stations maintained by TCEQ. The magnitude of the dots shows how many times the averaged daily PM_2.5_ concentration in a specific station exceeded the 35 μg/m^3^ standard established by EPA from 2000-2019**

**S4 Fig. Map showing the sum of reported environmental release events from 2000-2020 and leaking petroleum tanks within each census tract**

**S5 Fig. Map showing the second environmental variable, which is the sum of the number of dry cleaners, petroleum tanks, and sites that are part of an Industrial and Hazardous Waste Corrective Action (IHWCA) program defined by the TCEQ within each census tract**

**S6 Fig. Map showing flood vulnerability based on the inundation method**

**S7 Fig. Map showing the percentage of age 25 years and over to the total population in each census tract with a high school diploma or higher degree**

**S8 Fig. The averaged exceedance in the 5 categories for the tracts with the highest and lowest overall vulnerability (highest averaged exceedance in 5 categories) within Harris County**

**S9 Fig. Results of the K-means cluster analysis. Different colors reflect the belongingness of tracts to the three classes defined in the method for Category 1: Access to Medical (see Table 1 for more details). Areas with hatched lines represent census tracts with missing data on chronic disease risk factors, health outcomes, and clinical preventive service form [41]**

**S10 Fig. Results of the K-means cluster analysis. Different colors reflect the belongingness of tracts to the three classes defined in the method for Category 2: Underlying Conditions (see Table 1 for more details). Areas with hatched lines represent census tracts with missing data on chronic disease risk factors, health outcomes, and clinical preventive service form [41]**

**S11 Fig. Results of the K-means cluster analysis. Different colors reflect the belongingness of tracts to the three classes defined in the method for Category 3: Environmental Exposures (see Table 1 for more details)**

**S12 Fig. Results of the K-means cluster analysis. Different colors reflect the belongingness of tracts to the three classes defined in the method for Category 4: Vulnerability to Natural Disasters (see Table 1 for more details)**

**S13 Fig. Results of the K-means cluster analysis. Different colors reflect the belongingness of tracts to the three classes defined in the method for Category 5: Sociodemographic, Behavioral, and Lifestyle Factors (see Table 1 for more details). Areas with hatched lines represent census tracts with missing data on chronic disease risk factors, health outcomes, and clinical preventive service form [41]**

**S14 Fig. Results of the K-means analysis method. The graduated colors reflect the percentile of the overall vulnerability. Areas with hatched lines represent census tracts with missing data on chronic disease risk factors, health outcomes, and clinical preventive service form [41]**

**S15 Fig. Number of potential hospitalized COVID-19 patients based on age distribution and rates reported by the CDC [50]**

**S16 Fig. Number of potential hospitalized COVID-19 patients based on age distribution and rates reported by the New York Department of Health and Mental Hygiene [49]**

**S17 Fig. The mortality rate for each census tract based on age and sex distribution. Rates associated with each age interval were extracted from [52]**

**S1 Table. Rotated component matrix from the Principal Component Analysis. Varimax with Kaiser Normalization was chosen as the rotation method. Highlighted values show values greater than 0.5 or the maximum among the components**

**S2 Table. Descriptive summary statistics for all 46 variables. Median and median absolute deviation are reported instead of mean and standard deviation because none of the datasets were normally distributed**

**S3 Table: Results of the K-Means analysis: final cluster centers for different variables at each category**

## Notes

### Competing Interest Statement

The authors have declared no competing interest.

### Funding Statement

Funding provided by National Science Foundation grant #1759440 and grant # 1840607 and from the Hurricane Resilience Research Institute at the University of Houston.

## References

1. Johns Hopkins Centers for Civic Impact. Coronavirus resource center. 2020 [cited 21 Apr 2020]. Available: https://coronavirus.jhu.edu/

2. Adams ML, Katz DL, Grandpre J. Population based estimates of comorbidities affecting risk for complications from COVID-19 in the US. medRxiv. 2020; 2020.03.30.20043919. doi:10.1101/2020.03.30.20043919

3. Chin T, Kahn R, Li R, Chen JT, Krieger N, Buckee CO, et al. U.S. county-level characteristics to inform equitable COVID-19 response. medRxiv. 2020; 2020.04.08.20058248. doi:10.1101/2020.04.08.20058248

4. Garg S, Kim L, Whitaker M, O’Halloran A, Cummings C, Holstein R, et al. Hospitalization Rates and Characteristics of Patients Hospitalized with Laboratory-Confirmed Coronavirus Disease 2019 — COVID-NET, 14 States, March 1-30, 2020. MMWR Morb Mortal Wkly Rep. 2020;69: 458–464. doi:http://dx.doi.org/10.15585/mmwr.mm6915e3

5. Hossain MA. Is the spread of COVID-19 across countries influenced by environmental, economic and social factors? medRxiv. 2020; 2020.04.08.20058164. doi:10.1101/2020.04.08.20058164

6. Nayak A, Islam SJ, Mehta A, Ko Y-A, Patel SA, Goyal A, et al. Impact of Social Vulnerability on COVID-19 Incidence and Outcomes in the United States. medRxiv. 2020; 2020.04.10.20060962. doi:10.1101/2020.04.10.20060962

7. Petrilli CM, Jones SA, Yang J, Rajagopalan H, O’Donnell LF, Chernyak Y, et al. Factors associated with hospitalization and critical illness among 4,103 patients with COVID-19 disease in New York City. medRxiv. 2020; 2020.04.08.20057794. doi:10.1101/2020.04.08.20057794

8. Madjid M, Safavi-Naeini P, Solomon SD, Vardeny O. Potential Effects of Coronaviruses on the Cardiovascular System: A Review. JAMA Cardiol. 2020. doi: 10.1001/jamacardio.2020.1286

9. Goyal P, Choi JJ, Pinheiro LC, Schenck EJ, Chen R, Jabri A, et al. Clinical Characteristics of Covid-19 in New York City. N Engl J Med. 2020. doi:10.1056/NEJMc2010419

10. Lewnard JA, Liu VX, Jackson ML, Schmidt MA, Jewell BL, Flores JP, et al. Incidence, clinical outcomes, and transmission dynamics of hospitalized 2019 coronavirus disease among 9,596,321 individuals residing in California and Washington, United States: a prospective cohort study. medRxiv. 2020; 2020.04.12.20062943. doi:10.1101/2020.04.12.20062943

11. Parohan M, Yaghoubi S, Seraj A, Javanbakht MH, Sarraf P, Djalali M. Risk factors for mortality of adult inpatients with Coronavirus disease 2019 (COVID-19): a systematic review and meta-analysis of retrospective studies. medRxiv. 2020; 2020.04.09.20056291. doi:10.1101/2020.04.09.20056291

12. Ranjan R. Estimating the Final Epidemic Size for COVID-19 Outbreak using Improved Epidemiological Models. medRxiv. 2020; 2020.04.12.20061002. doi:10.1101/2020.04.12.20061002

13. Tahmasebi P, Shokri-Kuehni SMS, Sahimi M, Shokri N. How do environmental, economic and health factors influence regional vulnerability to COVID-19? medRxiv. 2020; 2020.04.09.20059659. doi:10.1101/2020.04.09.20059659

14. Stephanie Lamm. These areas of Harris County are at highest risk from coronavirus. Houston Chronicle. 20 Mar 2020. Available: https://www.houstonchronicle.com/news/houston-texas/houston/article/Virus-could-hit-Harris-unevenly-15147054.php

15. Conticini E, Frediani B, Caro D. Can atmospheric pollution be considered a co-factor in extremely high level of SARS-CoV-2 lethality in Northern Italy? Environ Pollut. 2020; 114465. doi:https://doi.org/10.1016/j.envpol.2020.114465

16. Wang J, Tang K, Feng K, Lv W. High Temperature and High Humidity Reduce the Transmission of COVID-19. 2020.

17. Currie J, Zivin JG, Mullins J, Neidell M. What Do We Know About Short-and Long-Term Effects of Early-Life Exposure to Pollution? Annu Rev Resour Econ. 2014;6: 217–247. doi:10.1146/annurev-resource-100913-012610

18. Erickson TB, Brooks J, Nilles EJ, Pham PN, Vinck P. Environmental health effects attributed to toxic and infectious agents following hurricanes, cyclones, flash floods and major hydrometeorological events. J Toxicol Environ Heal Part B. 2019;22: 157–171. doi:10.1080/10937404.2019.1654422

19. Landrigan PJ, Wright RO, Cordero JF, Eaton DL, Goldstein BD, Hennig B, et al. The NIEHS superfund research program: 25 years of translational research for public health. Environ Health Perspect. 2015;123: 909–918. doi:10.1289/ehp.1409247

20. Webber B, Stone R. Incidence of Non-Hodgkin Lymphoma and residential proximity to Superfund sites in Kentucky. J Environ Health. 2017;80: 22–29.

21. Bennett JE, Tamura-Wicks H, Parks RM, Burnett RT, Pope CA, Bechle MJ, et al. Particulate matter air pollution and national and county life expectancy loss in the USA: A spatiotemporal analysis. PLoS Med. 2019;16: e1002856. doi:10.1371/journal.pmed.1002856

22. Kiaghadi A S. Rifai H. Chemical, and microbial quality of floodwaters in Houston following Hurricane Harvey. Environ Sci & Technology. 2019;53: 4832–4840. doi:https://doi.org/10.1021/acs.est.9b00792

23. Marcantonio RA, Field S, Regan PM. Toxic trajectories under future climate conditions. PLoS One. 2019;14: e0226958. Available: https://doi.org/10.1371/journal.pone.0226958

24. Islam MS, Bonner JS, Fuller CS, Kirkey W. Impacts of an extreme weather-related Episodic event on the Hudson River and estuary. Environ Eng Sci. 2016;33: 270–282. doi:10.1089/ees.2015.0564

25. Patanavanich R, Glantz SA. Smoking is Associated with COVID-19 Progression: A Meta-Analysis. medRxiv. 2020; 2020.04.13.20063669. doi:10.1101/2020.04.13.20063669

26. CDC COVID-19 Response Team. Geographic Differences in COVID-19 Cases, Deaths, and Incidence — United States, February 12–April 7, 2020. MWR Morb Mortal Wkly Rep. 2020;69: 465–471. doi:http://dx.doi.org/10.15585/mmwr.mm6915e4

27. Nogee D, Tomassoni A. Concise Communication: Covid-19 and the N95 Respirator Shortage: Closing the Gap. Infect Control Hosp Epidemiol. 2020/04/13. 2020; 1–4. doi:DOI: 10.1017/ice.2020.124

28. Carenzo L, Costantini E, Greco M, Barra FL, Rendiniello V, Mainetti M, et al. Hospital surge capacity in a tertiary emergency referral centre during the COVID-19 outbreak in Italy. Anaesthesia. 2020;n/a. doi:10.1111/anae.15072

29. John Eligon, Burch ADS, Dionne Searcey, Jr. RAO. Black Americans Face Alarming Rates of Coronavirus Infection in Some States. The New York Times. 7 Apr 2020. doi:https://www.nytimes.com/2020/04/07/us/coronavirus-race.html

30. Nelson A. Unequal treatment: Confronting racial and ethnic disparities in health care. J Natl Med Assoc. 2002;94: 666–668. Available: http://search.proquest.com.ezproxy.lib.uh.edu/docview/214066442?accountid=7107

31. Institute of Medicine. Crossing the Quality Chasm: A New Health System for the 21st Century. Washington, DC: The National Academies Press; 2001. doi:10.17226/10027

32. Chetty R, Stepner M, Abraham S, Lin S, Scuderi B, Turner N, et al. The association between income and life expectancy in the United States, 2001-2014. JAMA - J Am Med Assoc. 2016. doi:10.1001/jama.2016.4226

33. U.S. Census. New Census Bureau Estimates Show Counties in South and West Lead Nation in Population Growth. In: United States Census Bureau [Internet]. 2019 [cited 20 Apr 2020]. Available: https://www.census.gov/newsroom/press-releases/2019/estimates-county-metro.html

34. Manson S, Schroeder J, Van Riper D, Ruggles S. IPUMS national historical geographic information system: Version 14.0 [Database]. 2019 [cited 10 Feb 2020] p. Minneapolis, MN. Available: http://doi.org/10.18128/D050.V14.0

35. U.S. Federal Emergency Management Administration (FEMA). FEMA - Harvey damage assessments and claims. 2018. doi:https://doi.org/10.4211/hs.73c4f3dcff884a6da2c0982df769987c

36. Abdulla B, Kiaghadi A, Rifai HS, Birgisson B. Characterization of vulnerability of road networks to fluvial flooding using SIS network diffusion model. J Infrastruct Preserv Resil. 2020;1: 6. doi:10.1186/s43065-020-00004-z

37. HRSA. Health Resources and Services Administration (HRSA) query data explorer tool. 2020 [cited 13 Apr 2020]. Available: https://data.hrsa.gov/tools/data-explorer

38. Harris County Health System. Harris health system locations. 2020 [cited 13 Apr 2020]. Available: https://www.harrishealth.org/locations/hhs

39. The Department of Homeland Security. Homeland Infrastructure Foundation-Level Data (HIFLD) - Urgent Care Facilities. 2018 [cited 12 Apr 2020]. Available: https://hifld-geoplatform.opendata.arcgis.com/datasets/urgent-care-facilities

40. Microsoft. Microsoft Bing Maps Platform APIs. 2020 [cited 14 Apr 2020]. Available: https://www.bingmapsportal.com/

41. HealthLanscape. The 500 Cities Mapper. 2020. Available: https://www.healthlandscape.org/500Cities/

42. Texas Commission on Environmental Quality. Texas Air Monitoring Information System (TAMIS) database. 2020 [cited 8 Apr 2020]. Available: https://www17.tceq.texas.gov/tamis/index.cfm

43. United States Coast Guard. National Response Center. 2020 [cited 12 Apr 2020]. Available: https://nrc.uscg.mil/

44. Texas Commission on Environmental Quality. TCEQ GIS data. 2020. Available: https://www.tceq.texas.gov/gis/download-tceq-gis-data

45. U.S. Environmental Protection Agency. Superfund: national priorities list (NPL). In: EPA [Internet]. 2019 [cited 2 Jan 2020]. Available: https://www.epa.gov/superfund/superfund-national-priorities-list-npl

46. Patil VH, Singh SN, Mishra S, Donavan DT. Parallel analysis engine to aid determining number of factors to retain [Computer software]. Available from http://smishra.faculty.ku.edu/parallelengine.htm. 2007.

47. Johns Hopkins University. COVID-19 Dashboard by the Center for Systems Science and Engineering (CSSE) at Johns Hopkins University (JHU). 2020 [cited 21 Apr 2020]. Available: https://coronavirus.jhu.edu/map.html

48. Texas Case Counts, COVID-19, Coronavirus Disease 2019. In: Texas Department of State Health and Services [Internet]. 2020 [cited 22 Apr 2020]. Available: https://txdshs.maps.arcgis.com/apps/opsdashboard/index.html#/ed483ecd702b4298ab01e8b9cafc8b83

49. New York City Department of Health and Mental Hygiene. COVID-19: Data. 2020 [cited 23 Apr 2020]. Available: https://www1.nyc.gov/site/doh/covid/covid-19-data.page

50. CDC COVID-19 Response Team. Laboratory-Confirmed COVID-19-Associated Hospitalizations. In: Centers for Disease Control and Prevention [Internet]. 2020 [cited 22 Apr 2020]. Available: https://gis.cdc.gov/grasp/COVIDNet/COVID19_3.html

51. Verity R, Okell LC, Dorigatti I, Winskill P, Whittaker C, Imai N, et al. Estimates of the severity of coronavirus disease 2019: a model-based analysis. Lancet Infect Dis. 2020. doi:10.1016/S1473-3099(20)30243-7

52. Flavia Riccardo, Xanthi Andrianou, Antonino Bella, Martina Del Manso AMU, Massimo Fabiani, Stefania Bellino, Stefano Boros, Fortunato (Paolo) D’Ancona MCR, Antonietta Filia, Ornella Punzo, Andrea Siddu, Corrado Di Benedetto, Marco Tallon AC, Maria Rita Castrucci, Patrizio Pezzotti, Paola Stefanelli GR. COVID-19 outbreak, National update: March 23, 2020 (in Italian). Rome, Italy; 2020. Available: https://www.epicentro.iss.it/coronavirus/bollettino/Bollettino-sorveglianza-integrata-COVID-19_26-marzo2020.pdf

53. Souri AH, Choi Y, Jeon W, Li X, Pan S, Diao L, et al. Constraining NOx emissions using satellite NO2 measurements during 2013 DISCOVER-AQ Texas campaign. Atmos Environ. 2016;131: 371–381. doi:https://doi.org/10.1016/j.atmosenv.2016.02.020

54. Souri AH, Choi Y, Li X, Kotsakis A, Jiang X. A 15-year climatology of wind pattern impacts on surface ozone in Houston, Texas. Atmos Res. 2016;174-175: 124–134. doi:https://doi.org/10.1016/j.atmosres.2016.02.007

55. Sillman S. The relation between ozone, NOx and hydrocarbons in urban and polluted rural environments. Atmos Environ. 1999;33: 1821–1845. doi:https://doi.org/10.1016/S1352-2310(98)00345-8

56. Sadeghi B, Choi Y, Yoon S, Flynn J, Kotsakis A, Lee S. The characterization of fine particulate matter downwind of Houston: Using integrated factor analysis to identify anthropogenic and natural sources. Environ Pollut. 2020;262: 114345. doi:https://doi.org/10.1016/j.envpol.2020.114345

57. Wu X, Nethery RC, Sabath BM, Braun D, Dominici F. Exposure to air pollution and COVID-19 mortality in the United States. medRxiv. 2020; 2020.04.05.20054502. doi:10.1101/2020.04.05.20054502

58. Opportunity Zones. In: City of Hosuton [Internet]. 2020 [cited 20 Apr 2020]. Available: https://www.houstontx.gov/opportunityzones/index.html

59. Yang W, Deng M, Li C, Huang J. Spatio-Temporal Patterns of the 2019-nCoV Epidemic at the County Level in Hubei Province, China. International Journal of Environmental Research and Public Health. 2020. doi:10.3390/ijerph17072563

60. Weiming T, Huipeng L, Gifty M, Zaisheng W, Weibin C, Dan W, et al. The changing patter of COVID-19 in China: A tempo-geographic analysis of the SARS-CoV-2 epidemic. Clin Infect Dis. 2020; ciaa423. doi:10.1093/cid/ciaa423

61. Flanagan B, Gregory E, Hallisey E, Heitgerd J, Lewis B. A Social Vulnerability Index for Disaster Management. J Homel Secur Emerg Manag. 2011;8: 22. doi:10.2202/1547-7355.1792

62. Butler DC, Petterson S, Phillips RL, Bazemore AW. Measures of Social Deprivation That Predict Health Care Access and Need within a Rational Area of Primary Care Service Delivery. Health Serv Res. 2013;48: 539–559. doi:10.1111/j.1475-6773.2012.01449.x

